# The self-management support needs of people newly diagnosed with psoriatic arthritis: a realist review protocol

**DOI:** 10.1101/2025.09.04.25335087

**Authors:** Katie Fishpool, Christine Silverthorne, Melanie Brooke, Leah Morris, Neil McHugh, Jason Ovens, Theresa Smith, William Tillett, Emma Dures

## Abstract

**Introduction:** Psoriatic arthritis is a form of inflammatory arthritis linked to psoriasis. Previous research from the UK has found that many people feel unsupported when newly diagnosed with psoriatic arthritis and lack confidence in managing their condition. This realist review aims to understand what works and does not work for whom and in what circumstances, in relation to healthcare professionals engaging with people to support them in developing self-management skills.

**Methods and analysis:** This protocol was developed by defining the scope of the review, using a brief directed literature review to support discussion by an expert group of researchers, healthcare professionals and a patient partner. A theoretical domains framework was generated, consisting of nine initial programme theories. These were further refined with input from Patient and Public Involvement and Engagement (PPIE) groups and used to develop a database search strategy.

A systematic database search will be carried out, supplemented by citation tracking, exploration of grey literature and a mixed methods survey of rheumatology health professionals. Data selection will be performed by a minimum of two reviewers and data from included sources will be extracted using a template. Data will be synthesised narratively with respect to the identified initial programme theories, using this data to refine or refute these theories. This will generate refined programme theories to explain what works for whom and in what circumstances.

**Ethics and dissemination:** Ethical approval for the health professionals survey was granted through the Research Ethics Committee, University of the West of England (Project ID: 10991848). Outputs will be disseminated to the research community through conference presentations and a peer-reviewed journal article. The strategy for sharing outputs with patients and health professionals will be discussed and agreed with knowledge user groups.

**Strengths and limitations of this study:**

- The use of realist methodology will facilitate exploration of the elements of self-management support that are beneficial, who they benefit and under what circumstances.
- PPIE contributions throughout the review process will ensure that the protocol, findings and subsequent discussion are relevant and reflect the lived experience of people with psoriatic arthritis.
- Only evidence available in English language will be included.

## INTRODUCTION

Psoriatic arthritis (PsA) is a form of inflammatory arthritis that is relatively unknown in the general population, affecting an estimated 190,000 adults in the UK (1). PsA is equally prevalent in men and women with a typical onset between the ages of 30 and 50 years, and main symptoms are pain, inflammation, stiffness and fatigue. Around 20% of people who have psoriasis will also develop PsA (2), with approximately 8,430 adults newly diagnosed with PsA each year in the UK(1).

Self-management involves the interplay of many tasks and skills required to focus on health needs, activate resources and live well with a chronic condition (3). It is a key strategy for reducing the disease burden on individuals, healthcare systems and society and there are national policies supporting the role of self-management as a tool to improve the health of the nation and reduce pressures on the National Health Service (NHS) (4). Supported self-management involves empowering patients to make decisions about their own health and to effectively utilise available healthcare resources, with the aim of achieving increased independence.

There are no specific guidelines for supporting self-management of PsA but there are recommendations from the European Alliance of Associations for Rheumatology (EULAR) on the implementation of self-management strategies in patients with inflammatory arthritis (IA) including PsA. These recommendations emphasise that self-management is a multicomponent and complex intervention which should be supported by healthcare professionals, however, it is often an unmet need in those with IA (5).

The experience of people newly diagnosed with PsA may differ from other forms of IA. There can be significant delays in the diagnosis of PsA, with many patients experiencing symptoms for several years before receiving a formal diagnosis and starting treatment (6). This delay can lead to worse treatment outcomes and has a detrimental impact on the health and wellbeing of individual patients. Screening and education about PsA is recommended for people diagnosed with psoriasis (7) but this recommendation is often neglected despite evidence that screening has positive benefits for patients (8).

The treatment outcomes which are prioritised by people with PsA have been previously identified (9). Diagnosis was often viewed as a turning point, bringing feelings of validation and relief but also anger and sadness about the impact of the delays (10). People highlighted the importance of self-management and doing what was possible to keep themselves well to minimise the impact of PsA, but they required healthcare professionals to understand their priorities and access to resources to successfully support this.

### Review aim and objectives

The aim of the review is to understand how existing self-management resources work, including for whom and in which contexts. The objectives of the review are to:

1. Develop a programme theory of how existing self-management resources support patients newly diagnosed with PsA, for whom and in what circumstances.
2. Identify a research agenda to address gaps that emerge from the review, including patient groups with unmet needs.

## METHOD AND ANALYSIS

A realist approach to evidence synthesis acknowledges that interventions may work in some contexts but not others. This is often represented as:

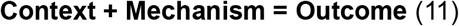

For this review, the **outcome** of interest is the development of a person’s ability to self-manage their PsA. The **mechanisms** that result in self-management are often hidden and are influenced by the **context** in which they occur. This notion of outcomes being context-bound is a key principle of the realist approach (12). Developing theories about how the mechanisms that result in self-management are influenced by context, and testing these using the selected evidence will generate information about the impact of individual intervention components. This will enable knowledge users, such as health professionals, PPIE and researchers, to design and implement interventions that include only the components that are effective for their specific context (13).

The process for developing the review protocol and conducting the review are shown in Figure 1. Although displayed as a flow diagram, there is an expectation that some steps will be overlapping and iterative. As the aim of realist review is to refine theories, new evidence that emerges from the literature search or from questions raised by knowledge users can and should be included at any point (12).

**Figure 1.**
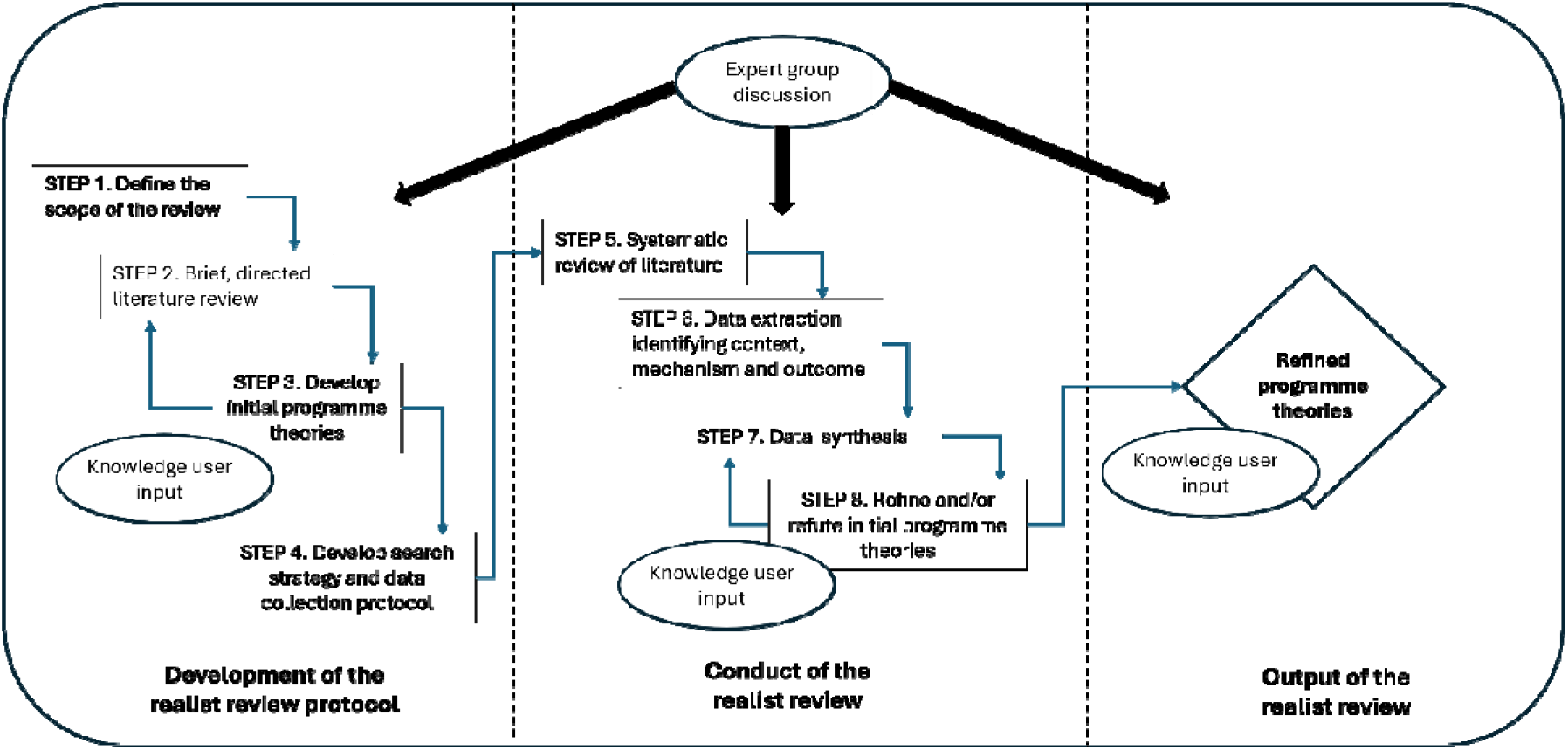
Realist review process diagram

**Figure 2.**
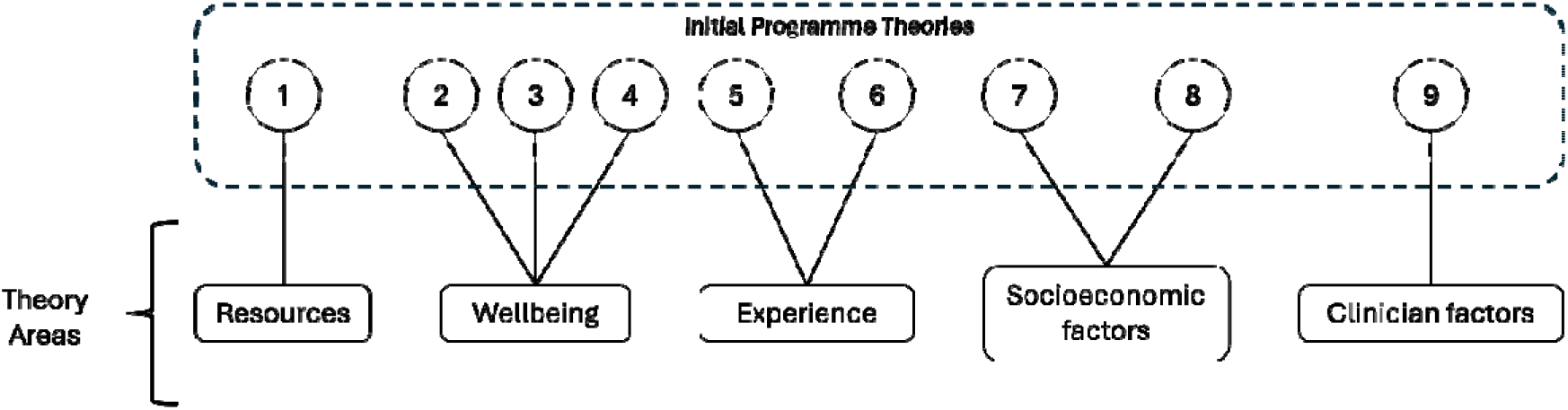
Theoretical domain framework

A survey of rheumatology health professionals is being conducted concurrently to complement the realist review (Appendix 2). This survey aims to discover what services are routinely offered to patients who are diagnosed with PsA and the opinions of health professionals on the suitability of these services.

### Step 1. Define the review scope

A person in the UK with suspected PsA will be referred to a rheumatology service for formal diagnosis. Their initial care following diagnosis will depend on disease severity but is likely to include clinical assessment, information sharing, treatment planning, symptom management and advice seeking opportunities with a rheumatology consultant or registrar, specialist nurses and other allied health professionals (AHPs) (14).

The programme architecture that is being investigated is the provision of support for self-management by rheumatology services for people in this stage of recent diagnosis. This review will explore the evidence for the mechanisms which support people to self-manage and the contexts which enable or impede these mechanisms.

### Step 2. Brief directed literature search

A brief literature search of MEDLINE, CINAHL Plus, EMBASE and APA PsycINFO was conducted by KF using the search terms “psoriatic arthritis” and “self-management”. This gave an impression of the type and volume of evidence available and was used to inform discussions about the development of the protocol and to generate the initial programme theories.

### Step 3. Develop initial programme theories

Initial programme theories (IPTs) articulate how programme components work to achieve outcomes (12) Ideas were formulated by an iterative process of creative brainstorming by the expert team, which consists of healthcare professionals and researchers with a range of backgrounds including nursing (KF), health psychology (CS and ED), clinical rheumatology (NJM and WT), a specialist subject librarian (JO) and a patient partner with lived experience of PsA (MB). This process was supported by a realist methodology specialist (LM) and the resulting IPTs were grouped under five theory areas in a theoretical domain framework. These IPTs are not intended to be definitive or conclusive, but to provide an informed starting point for investigating the available evidence.

Knowledge user workshops and interviews were held to discuss the IPTs. They were open to adults with a diagnosis of PsA and advertised through national patient organisation websites. Fourteen people with lived experience of PsA reviewed the proposed IPTs and gave feedback on these, providing a robust basis for developing and prioritising the theories that will be tested by the review (15).

Theory Area 1: Resources offered – the content of self-management resources offered will impact on how patients engage with self-management.

Initial programme theory

1. IF the resources offered are not condition specific THEN people who have recently been diagnosed with PsA may not engage with self-management support BECAUSE they cannot understand how it is relevant to them. Theory Area 2: Wellbeing – the physical and psychological wellbeing of a person newly diagnosed with PsA impacts on their ability to engage with self-management support. Initial programme theories
2. IF PsA symptoms (skin involvement, unpredictable joint pain) are not addressed THEN self-management advice will be difficult to implement BECAUSE it feels unachievable.
3. IF a person has an emotional support network THEN they will find it easier to engage with self-management BECAUSE they will be able to share and discuss their diagnosis, plans and concerns.
4. IF a person has a practical support network THEN they will find it easier to engage with self-management BECAUSE they will have help to adopt self-management behaviours. Theory Area 3: Patient experience – previous experiences of health and healthcare will impact on a person’s engagement with self-management of their PsA. Initial programme theories
5. IF a person had previous experience of self-managing psoriasis THEN they will be more able to manage their PsA BECAUSE they will have increased health literacy and familiarity with self-management practices.
6. IF a person has previous experience of self-managing psoriasis THEN they will be less able to manage their PsA BECAUSE they are already managing the burden of another condition(s). Theory Area 4: Socioeconomic factors – factors which may have an impact on a person’s ability to engage with self-management of their PsA. Initial programme theories
7. If a person is in employment THEN they will find self-management more difficult BECAUSE of the competing demands on their time and resources.
8. IF a person has caring responsibilities THEN they will find self-management of their PsA more difficult BECAUSE their ability to adapt their lifestyle and activities is reduced. Theory Area 5: Health professional factors – the knowledge, experience and attitude of healthcare professionals will influence their ability to engage people in self-management.
9. IF the care pathway for patients with PsA is not understood by health professionals THEN opportunities to offer support with self-management may be missed BECAUSE health professionals will not have the confidence or expertise to recognise and address these.

IPT5 and IPT6 are “rival theories” – theories which directly contradict each other (15). The evidence for the review will be analysed to understand the context in which one or the other theory may be correct.

The findings of the literature search will be analysed with regards to whether they support or refute the IPTs and, from this, refined context-mechanism-outcome (CMO) statements will be developed.

The full list of initial theories generated is shown in Appendix 1.

### Step 4. Develop search strategy and data collection protocols

The following electronic databases will be searched by a specialist subject librarian (JO) for evidence published in English language from January 2010 onwards; CINAHL, EMBASE, Emcare, MEDLINE, PsychINFO. There will be no restriction on the age of participants, allowing inclusion of evidence for people across the lifespan and highlighting any gaps in provision. A further search will be conducted for evidence from patient organisations that support people with PsA and the reference lists of included items will be hand searched to check for relevant papers not captured by the database searching.

### Step 5. Selection and appraisal of evidence

A minimum of two independent reviewers will screen the titles and abstracts (where applicable) of all discovered evidence against the stated inclusion and exclusion criteria (Table 1). Regular meetings will be held by KF, CS, ED, LM and MB throughout the title and abstract screening process to aid understanding and reduce disagreements (16). Papers that proceed to the full text stage of screening will also be reviewed by two or more independent reviewers who will document the main reason any excluded papers do not meet the inclusion criteria. Any differences in opinion between the reviewers will be resolved by discussing the papers, with an additional independent reviewer to support mediation, as required.

**Table 1.** Review inclusion and exclusion criteria.

| Inclusion criteria |  |
| --- | --- |
| P - Population | People who have been diagnosed with PsA within the last 24 months<br>Healthcare professionals involved in the treatment of people who have recently been diagnosed with PsA |
| I - Intervention | Resources, methods, tools or interventions that support and promote self-management of PsA |
| C- comparator | None |
| O - outcomes | Adjustments or acceptance of self-management, shared decision making, confidence to manage health<br>Knowledge, attitude or skills of healthcare professionals |
| Study design |  |
| No restrictions on study design<br>Include grey literature through citation searches and identification by the review team |  |
| Exclusion criteria |  |
| Data related to PsA is not clearly stratified<br>Evidence that is not available in English |  |

### Step 6. Data extraction

We will develop and pilot a bespoke data extraction form to answer our review aim. The extracted data will be mapped onto the IPTs within the theoretical domains’ framework, with an additional column to allow for any data which addresses the review aim but that has not been covered in the IPTs. Data from the included evidence will be extracted by two reviewers (KF and CS) and a third reviewer (ED) will extract a sample of evidence to ensure integrity of extraction. The use of a template will standardise data extraction and ensure that included evidence is interrogated across all identified theory areas (17). Any disagreements that may arise between the reviewers will be resolved by consensus, with an additional reviewer to support mediation. If necessary, the authors of the included papers will be contacted for further information or data clarification.

### Step 7. Data synthesis

Narrative reporting methods will be utilised due to the inclusion of qualitative data and overall heterogeneity of evidence sources included in this review. The findings from different studies will be compared to the IPTs, seeking both confirmatory and contradictory findings in order to understand alternative contexts and mechanisms. The synthesis will be guided by the theoretical domain framework to explore the aims of the review and identify gaps in the evidence.

### Step 8. Refine or refute initial programme theories

The synthesised data will be discussed by the expert group and with PPIE contributors to identify the areas of initial theory which are supported by the evidence, those which are not, and any new areas of theory that may have emerged during the review process. These refined programme theories will be the output of the realist review.

### Patient and public involvement statement

MB is a patient partner and has been involved in the review from inception, as a co-applicant on the funding supporting this review and contributing to each stage of the protocol development. She has also supported the review team to reach underserved populations for participation in PPIE workshops, specifically male participants. MB will be part of the team which analyses the extracted data and prepares final outputs from the review.

PPIE workshops and interviews have been arranged pragmatically to discuss experiences of diagnosis and management of PsA. Discussions were analysed for themes and key words which contributed to development of the initial programme theories. A strong sense of lack of support for self-management was apparent across all PPIE activity, validating the decision to investigate this gap in healthcare provision. A list of the IPTs was shared with everyone who had attended a PPIE workshop or interview at this stage and 14 people provided feedback which was used to further refine the IPTs.

Future workshops and interviews are planned to refine and test the theories developed from the review data. The inclusion and outcome of all PPIE activities will be reported in the final review manuscript using the GRIPP2 short-form reporting checklist (18), which is a tool specifically designed to improve the reporting of PPIE in research.

## Data Availability

All data produced in the present study are available upon reasonable request to the authors

## ETHICS AND DISSEMINATION

Ethical approval for the health professional survey was granted through the Research Ethics Committee, University of the West of England (Project ID: 10991848). The findings of this review will be disseminated via relevant peer-reviewed journals, conference presentations and through sharing findings with relevant charities and health professionals.

## AUTHOR CONTRIBUTIONS

Project funding application by ED and NM with support from WT, TS and MB. Review search strategy by KF, CS, JO and ED, database searches carried out by JO. PPIE activities arranged and facilitated by CS, with support from MB. Drafting of protocol by KF in discussion with CS, ED, LM and MB. Manuscript reviewed by all authors.

