## Appendix 1 for "The self-management support needs of people newly diagnosed with psoriatic arthritis: a realist review protocol"

| **PSA SPECIFIC FACTORS** | **COMMUNICATION OF SUPPORT** | **PSYCHOLOGICAL WELLBEING** | **EXPERIENCE OF HEALTHCARE** | **HOME LIFE** | **FINANCIAL CONCERNS** | **SERVICES** | **CLINICIANS** |
| --- | --- | --- | --- | --- | --- | --- | --- |
| IF a person has skin involvement in their PsA THEN it may have a greater impact on their psychological wellbeing (*and so may find It harder to engage with self-management support*) | IF a person has limited understanding of what self-management means THEN they may not think it is applicable to them | IF a person is feeling depressed about their diagnosis THEN they will find it difficult to manage their PsA diagnosis | IF a person has had a previous bad experience with medical staff THEN they will find it harder to engage with self-management support. | IF someone has lots of 'other' (non-health related) challenges going on in their daily lives, THEN self-management is less likely to be a personal priority. | IF a person has a supportive Employer THEN it will be easier for them to attend self-management support sessions | IF clinicians have limited time with patients THEN self-management support will be limited | IF clinicians have limited time with patients THEN self-management support will be limited |
| IF a person has not previously heard of PsA THEN they may not understand how their condition can be managed | IF the self-management information offered to patients bears no relation to their experiences so far THEN it will not feel pertinent/relevant | IF a person is finding it difficult To accept their PsA diagnosis THEN they will find it difficult to accept self-management support. | IF a person has previous experience of managing a long term condition THEN they will be more able to manage their diagnosis of PsA | IF a person has a supportive partner/family THEN they will find it easier to engage with self-management support. | IF a person is in a full-time job THEN they will find it harder to manage their diagnosis of PsA | IF a clinician believes that another member of the team is responsible for discussing self-management THEN they may not address it during their consultation | IF health professionals have pre-conceived ideas about which patients have (or will acquire) the skills, knowledge and confidence to self-manage THEN it will influence the help and support that they offer to different patients. |
| IF the support offered is not condition specific THEN people may not understand how it will help | IF the self-management help/advice/guidance that patients are offered does not align with their values, beliefs and practices THEN they are less likely to try it out. | IF a person is overwhelmed by their diagnosis THEN they may not be able to process information about self- management | IF a person has waited a very long time for a diagnosis THEN they may have less trust in health services and healthcare professionals | IF a person has young children or other caring responsibilities THEN they will find self-management of their PsA more difficult | IF someone has limited time and/or access to resources (e.g., physical activity classes and nutritious food) THEN they might struggle to self-manage their PsA | IF the care pathway for newly diagnosed patients is not understood by clinicians THEN opportunities to offer support with self management may be missed | IF a clinician is not confident in addressing self-management THEN they may not do so |
| IF rheumatology staff are not knowledgeable about psoriasis THEN they may find it difficult to give self-management advice | IF a person does not have English as their first language THEN they will find it harder to access self-management support | IF a person finds it difficult to talk to others | IF a person already has a long-term health condition when they are diagnosed with PsA THEN they will find it harder to manage their PsA as well | IF a person is in a full-time job THEN they will find it harder to manage their diagnosis of PsA |  | IF a person has waited a very long time for a diagnosis THEN they may have less trust in health services and healthcare professionals | IF rheumatology staff are not knowledgeable about psoriasis THEN they may find it difficult to give self-management advice |
|  |  | about their diagnosis THEN they will find it harder to follow self-management guidance. |  |  |  |  |  |
|  | IF someone does not have access to understandable information THEN they will find it harder to self-manage their health/PsA | IF someone thinks that there is nothing they can do to change their health (fatalistic) THEN they are less likely to change/adapt their behaviours. | IF people feel that their concerns about medication have not been addressed THEN they are less likely to engage with their healthcare providers | IF someone has limited time and/or access to resources (e.g., physical activity classes and nutritious food) THEN they might struggle to self-manage their PsA |  |  | IF the care pathway for newly diagnosed patients is not understood by clinicians THEN opportunities to offer support with self management may be missed |
|  | IF information is communicated by a professional that the patient holds in high regard THEN they are more likely to engage with advice | IF a person has skin involvement in their PsA THEN it may have a greater impact on their psychological wellbeing (*and so may find It harder to engage with self-management support*) | IF a person has limited understanding of what self-management means THEN they may not think it is applicable to them |  |  |  | IF people feel that their concerns about medication have not been addressed THEN they are less likely to engage with their healthcare providers |
|  |  |  | IF information is communicated by a professional that the patient holds in high regard THEN they are more likely to engage with advice |  |  |  |  |
