## Appendix 2 for "The self-management support needs of people newly diagnosed with psoriatic arthritis: a realist review protocol"

**Online Survey**
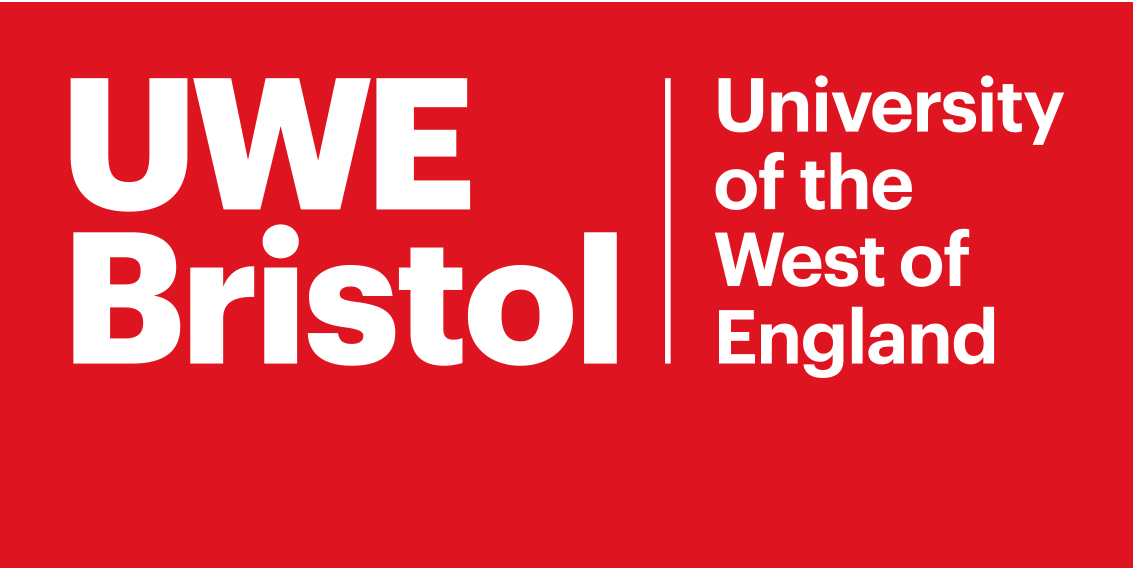

**Welcome to this brief survey about** **how helpful self-management resources are in meeting the support needs of patients newly diagnosed with Psoriatic Arthritis (PsA).**

We are a group of researchers led by Professors Emma Dures and Neil McHugh and funded by NIHR.

We would like to find out about:

1. The self-management support available to patients in your rheumatology team within the first 2 years of being diagnosed with psoriatic arthritis (PsA). This could include providing help and education around medications, acceptance and mental wellbeing, communicating to others about PsA and dealing with fatigue.

2. Whether you feel there are any gaps in the self-management support provided and your thoughts on how this could be improved.

3. Whether you feel there are any specific groups of patients who are not receiving the self-management support available and your thoughts on how we could better reach them.

**You need to be a rheumatology health professional working with patients with psoriatic arthritis to complete this survey.**

By filling out this survey, we presume you are providing your consent. If you would like to read the full details about the survey, please click here Participant Information Sheet

All information provided will be kept in adherence with GDPR. Privacy notice for research participants

If appropriate, please forward the link to this survey to rheumatology colleagues working in other teams.

Please click the 'Next page' button to begin.

**You have been invited to complete this survey as we want to understand how helpful self-management resources are in meeting the support needs of patients newly diagnosed with psoriatic arthritis (PsA).**

1) Which NHS Trust is your rheumatology team in?

TEXT BOX:

2) What do you think the main self-management support needs are for patients newly diagnosed with PsA (within the last 2 years)? **Please tick all that apply:**

- Questions about treatments / medication
- Questions about psoriasis
- A need for emotional support / help with acceptance and mental wellbeing
- Work
- Relationships / communicating to others about PsA
- Dealing with fatigue
- Other

TEXT BOX:

3) Do you think there are self-management support needs that are specific to patients with PsA?

- Yes (please describe below)
- No

TEXT BOX:

4a) Are patients newly diagnosed with PsA routinely given self-management support in your rheumatology team?

- Yes
- No
- Sometimes

4b) If self-management support is given, how is this done? **Please tick all that apply:**

- Group education/information session(s).
- Group self-management session(s).
- One-to-one support.
- Referral/signpost to external source of help.
- Provide self-management literature.
- Other.

TEXT BOX:

4c) If self-management support is given, who provides this support?

TEXT BOX:

5) Do you/your rheumatology team work with your hospital’s Dermatology department?

- Yes
- No
- Sometimes

6a) Do you/your rheumatology team refer and/or signpost patients to self-management support from an external provider?

- Yes
- No
- Sometimes

6b) Which external providers are patients referred and/or signposted to?

TEXT BOX:

6c) Who funds this support?

TEXT BOX:

7) Are there groups of patients with PsA who are **more** likely to access self-management support in your rheumatology service and why might this be?

- Yes (please describe below)
- No

TEXT BOX:

8) Are there groups of patients with PsA who are **less** likely to access self-management support in your rheumatology service and why might this be?

- Yes (please describe below)
- No

TEXT BOX:

9) Do you feel there are any gaps / areas of unmet self-management support needs?

- Yes (please describe below)
- No

TEXT BOX:

10) Do you feel any additional resources or adaptations to existing resources are required?

- Yes (please describe below)
- No

TEXT BOX:

11) What are the main **barriers** to meeting the self-management support needs of patients newly diagnosed with PsA? **Please tick all that apply:**

- The cost of delivering self-management support.
- Not having dedicated staff to deliver self-management support.
- Not having enough time within clinical appointments to provide self-management support.
- Self-management support not being routinely addressed in rheumatology appointments.
- Other.

TEXT BOX:

12) What are the main **enablers** to meeting the self-management support needs of patients newly diagnosed with PsA? **Please tick all that apply:**

- Patients understanding what self-management support involves and where they can get this.
- Options for different modes / types of delivery (e.g. in-person or online; one-to-one or groups).
- Options for different types of support (e.g. medication, acceptance and wellbeing, communicating to others about PsA, fatigue)
- Self-management support provided to specific groups (e.g. age, gender, ethnicity) to ensure patients can identify with the group.
- Having self-management resource materials available to give to patients.
- Having self-management resource materials available in different languages.
- Incorporating self-management support into routine rheumatology appointments.
- Other.

TEXT BOX:

13) What do you think is the best way of implementing self-management support into routine practice?

TEXT BOX:

14) Overall, how do you rate the self-management support that your service is able to offer to patients?

**Excellent	Good	Adequate	Poor	Very poor**

15) Is there any other information that you would like to tell us about self-management support for PsA?

TEXT BOX:
